# Screening for anxiety and depression: Validation of the PHQ-4 in a large sample of rehabilitation patients

**DOI:** 10.1101/2025.10.13.25337934

**Authors:** Stefan Gschwenter, Dominik Garber, Benedikt Steininger

**Author notes:** corresponding author: Stefan Gschwenter, Pensionsversicherung, Friedrich-Hillegeist-Straße 1, 1021 Vienna, Austria. These authors contributed equally to this work (Stefan Gschwenter, Dominik Garber).

## Abstract

**Background:** Anxiety and depression are the most prevalent mental disorders in both general and rehabilitation populations. The Patient Health Questionnaire for Depression and Anxiety (PHQ-4) is an ultra brief screening instrument previously validated for use in general and primary care populations. We aimed to validate the PHQ-4 for the rehabilitation setting by establishing its factorial and convergent validity. The availability of such screenings for rehabilitation is critical, as psychological distress is an important contributor to rehabilitation outcomes.

**Methods:** The study sample consisted of 66,412 rehabilitation patients from 15 inpatient rehabilitation centers in Austria, covering rehabilitation of various (non-psychiatric) medical indications. To validate the PHQ-4’s factorial validity, we conducted confirmatory factor analyses (CFA) comparing one-factor (psychological distress) or two-factor (anxiety and depression) models. Multi-group CFAs assessed measurement invariance across age, gender, and medical indication. Convergent validity was tested using ordinal logistic regression to model the PHQ-4’s association with measures of quality of life, vocational impairment, and work ability. We conducted additional analyses to rule out spurious results based on the very large same size used.

**Results:** The results demonstrated an overall excellent fit for the two-factor solution, outperforming the one-factor solution. Reliability indices for the two-factor solution were adequate for a screening instrument. Measurement invariance was demonstrated across age, gender, and all but one tested medical indications. Ordinal logistic regression supported convergent validity, showing associations with age and common patient reported outcomes.

**Conclusions:** This study demonstrates that the PHQ-4 is a valid and reliable tool for screening for anxiety and depression in rehabilitation settings. Based on a substantial sample and preregistered analysis strategy, the findings will likely generalize to rehabilitation populations in other countries.

**Trial Registration:** The current study was approved by the Ethics Committee of the Medical University of Vienna ([MASKED FOR REVIEW: reference number]) and preregistered on the Open Science Framework ([MASKED LINK FOR REVIEW: https://osf.io/79fgb/?view_only=de7954e85040422db0db08d24ac7ed79]).

---

Anxiety and depression are the most prevalent mental disorders in the general population [1] and frequently co-occur in rehabilitation settings, including musculoskeletal conditions [2], chronic somatic diseases [3], and cancer [4]. Due to their prevalence, developing and validating effective screening instruments are of substantial clinical and public health importance. Previously, the depression screening instrument PHQ-2 and the anxiety disorder screening instrument GAD-2 were combined to form the Patient Health Questionnaire for Depression and Anxiety, PHQ-4 [5]. The PHQ-4 has been validated in both general populations and specific primary care populations through existing self-report anxiety and depression scales, clinical interviews, analysis of factorial validity, and analysis of divergent validity [6].

The PHQ-4’s practical advantages lie in its efficiency and short nature, which minimizes patient burden while maintaining psychometric integrity [7]. This makes it well-suited for clinical settings where time constraints often limit comprehensive psychological assessments. Compared to longer instruments such as the PHQ-9 or GAD-7, the PHQ-4 allows relatively rapid screening for patients requiring further evaluation, meeting the psychometric criteria of economy and acceptability [8]. However, it is essential to acknowledge the trade-off between concise instrument length and diagnostic precision, particularly in contexts where more detailed mental health assessments are deemed necessary.

Previous research highlighted the significance of psychological distress in rehabilitation patients. Brünger and Spyra [9] found that the prevalence of depressive symptoms in rehabilitation patients is approximately four times higher than in the general population, while in certain medical indications, more than one-third of patients exhibit comorbid depressive symptoms. Furthermore, anxiety and depression negatively affect treatment adherence and rehabilitation outcomes in a variety of medical indications [10–15].

Appropriate identification and treatment of these mental health comorbidities may improve rehabilitation outcomes, as they can otherwise impair recovery [16]. Therefore, it has been suggested that rehabilitation is an appropriate opportunity to screen for and initiate treatment for affective disorders [17].

Despite its proven utility in general populations [18] and primary care settings [19–21], the PHQ-4 has, to our knowledge, never been validated in a rehabilitation context. Furthermore, construct validation is a dynamic and context-dependent process that requires ongoing reassessment, especially when instruments are used across different populations or settings [22]. Even screening tools with well-established psychometric properties, such as the PHQ-4, may produce different results depending on the clinical setting in which they are applied. Consequently, in this study we assess the PHQ-4’s factorial and convergent validity through Confirmatory Factor Analysis (CFA) and examine its relationship with theoretically implicated health metrics, such as functional status, impairment and work ability. Our research is based on an extensive sample of over 66,000 rehabilitation patients in Austria, ensuring statistically robust results that are likely to be generalizable to rehabilitation populations in other countries. This is particularly relevant for settings where the PHQ-4 has already been validated for the general population, such as Austria [23].

## Method

This study’s sampling approach, used variables, hypothesis, and planned analyses were preregistered on Open Science Framework ([MASKED LINK FOR REVIEW: https://osf.io/79fgb/?view_only=de7954e85040422db0db08d24ac7ed79]) before any analyses were conducted. Deviations from our preregistered analyses can be obtained from Supplement D.

### Participants and Study Design

The present study is a multicenter, retrospective analysis and was approved by the Ethics Committee of the Medical University of Vienna ([MASKED FOR REVIEW: reference number]). Data were collected as part of the standard procedure during regular operations across 15 inpatient rehabilitation centers operated by [MASKED FOR REVIEW: health care provider]. The [health care provider] represents part of the public social insurance and healthcare system in Austria. The rehabilitation centers offer multidisciplinary rehabilitation programs for various medical indications. Patients attended inpatient rehabilitation for 3 to 4 weeks for therapeutic procedures to maintain and restore work capacity and participation. The PHQ-4, along with other instruments, was used as a screening tool at the beginning of each patient’s respective therapeutic process. All instruments were administered in German. Patient data from these centers were included for analysis from January 1, 2021, through November 1, 2023. Only patients with available PHQ-4 scores were included in the sample. The minimum age for inclusion in the study was 18 years, yielding data for 66,412 patients. Access date of patient data was December 1, 2023. Authors did not have access to information that could identify individual participants of this study.

### Measures

Demographic data, including age and gender, were collected at the rehabilitation centers as part of routine practice. These variables were recorded alongside other assessments, which were used to evaluate various psychological and health-related factors. Participants’ gender

### Patient Health Questionnaire-4 (PHQ-4)

The PHQ-4 [5] is an ultra-brief self-report questionnaire screening for depression and anxiety. It consists of two 2-item subscales: the PHQ-2 for depression and the GAD-2 for anxiety. These four questions are answered on a four-point scale ranging from 0 (not at all) to 3 (almost every day) in a self-report format that measure the severity of symptoms over the past two weeks. Validation studies have shown that it can be used as a two-dimensional instrument (for both anxiety and depression scores) and for creating an overall score. To calculate these scores, the responses to the individual items are summed. Thus, the subscale scores can range from 0 to 6, while the total score can range from 0 to 12. Higher values indicate greater psychological distress overall [18]. See Table 1 for a complete list of items in both English and German.

**Table 1.**
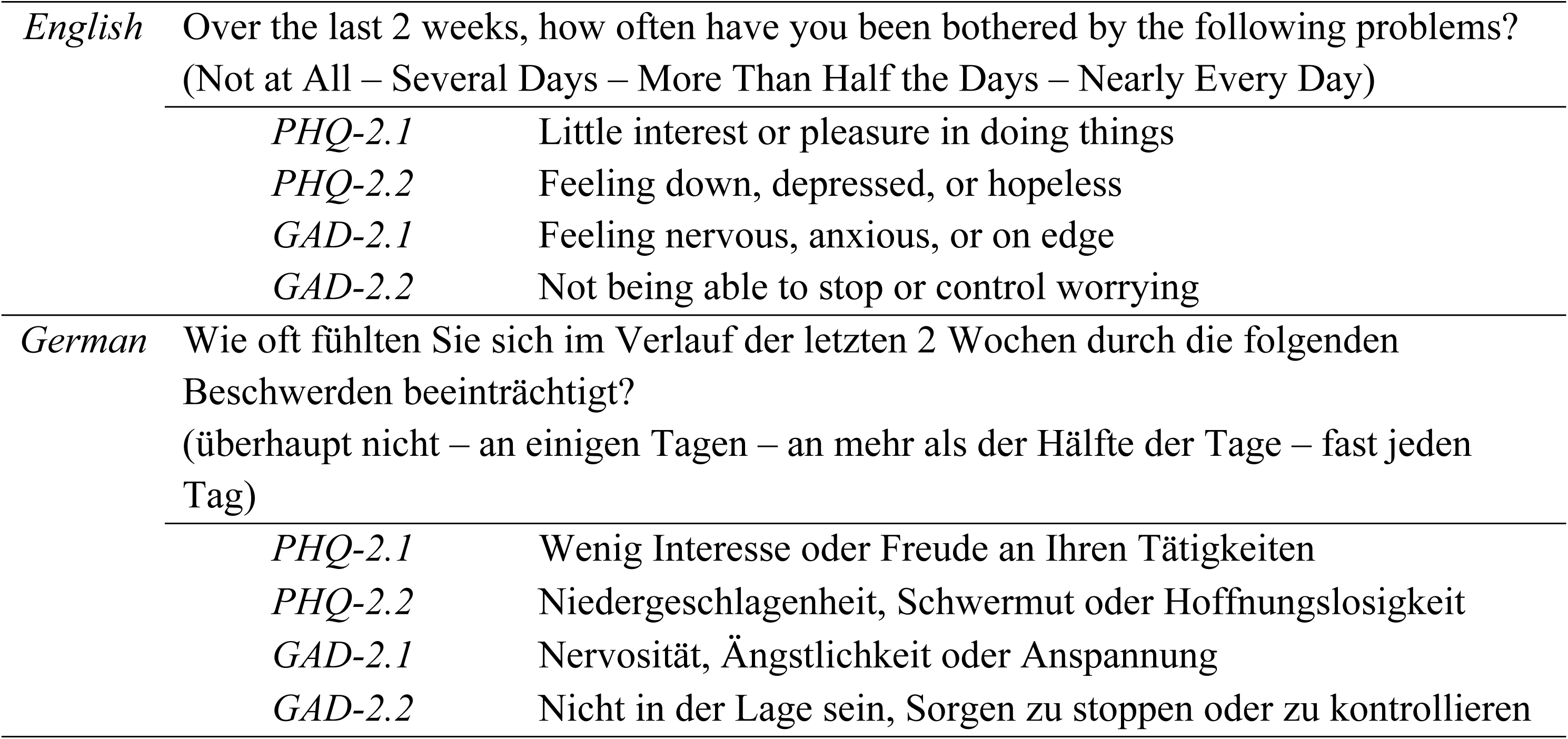
Items of the PHQ-4.

### European Quality of Life 5 Dimensions 5 Level Version (EQ-5D-5L)

The EQ-5D-5L [24] is a widely used instrument for measuring patient-reported outcomes, specifically focusing on subjective health-related quality of life (HRQOL). It evaluates five dimensions: mobility, self-care, usual activities, pain/physical discomfort and anxiety/depression. Each dimension is rated on a five-level scale ranging from 1 (no problems) to (5 extreme problems). This structure allows the EQ-5D-5L to define 3,125 unique health states, which can be converted into an EQ-5D-5L index value based on a set of preference values from a general population. As no specific value set for the Austrian population exists, the German value set was used for calculations [25]. It establishes an index score within the range of -.661 (extreme problems in all five dimensions) and 1 (no problems in any dimension). In addition to the index score, the instrument incorporates a visual analogue scale (VAS), wherein respondents rate their overall health on a scale ranging from 0 (worst imaginable health) to 100 (best imaginable health).

### Work Ability Index (WAI)

The WAI [26] is a self-report questionnaire used to assess an individual’s perceived work ability in relation to job demands, overall health status, and mental resources. The questionnaire consists of seven items, incorporating both numerical and categorical response formats, capturing multiple dimensions of work ability. A total WAI score is calculated by summing all item responses [27].

### Screening Instrument for the Identification of Need for Medical-Vocational Oriented Rehabilitation in Chronic Diseases (SIMBO-C)

The SIMBO-C [28] is a screening instrument developed to assess specific vocational problems in individuals with chronic diseases. It comprises of seven weighted criteria of impairment in vocational participation, designed to identify patients with extensive work-related complications. For the present study, only the item "To what extent does your current health condition affect your work?" was used. The item is answered on an 11-point scale, with higher values indicating greater impairment.

### Statistical Analysis

All statistical analyses were performed using R version 4.2.0 [29].

### Item Characteristics and Internal Consistency

We assessed item characteristics for the PHQ-4, including item, subscore and composite score means and standard deviations, as well as item-item correlations and item-total correlations. Where not specified otherwise, correlations are polychoric correlations, in line with the type of correlation used in our Confirmatory Factor Analysis estimation method (see next section). To assess reliability, we computed Mc Donald’s ω as a measure of internal consistency.

### Confirmatory factor analysis

To assess the factorial structure of the PHQ-4 we conducted confirmatory factor analyses (CFA), using the lavaan R package version 0.6-20.2330 [30]. We compared two main models: a one-factor structure in which all four items are subsumed under one factor, and a two-factor structure in which the two anxiety items loaded on an anxiety factor and the two depression items on a depression factor.

As the PHQ-4 items have an ordinal level of measurement and are usually not normally distributed [18], we opted not to use the common maximum likelihood estimator but instead weighted least-squares with mean and variance adjustment [WLSMV; 31, 32]. This estimation approach is well suited for fitting categorical, non-normal data [33]. Factor expected values were set to 0. Theta parameterization was employed in order to enable the planned analysis of measurement invariance.

Our interpretation of the results focuses on fit indices, i.e., the Root Mean Square Error of Approximation (RMSEA), Standardized Root Mean Square Residual (SRMR), Comparative Fit Index (CFI), and Tucker–Lewis index (TLI) for each model. RMSEA and CFI were subject to a correction suggested for the use with ordinal or categorical data in robust CFA methods [34]. This prevents a previously reported inflation of such fit indices when used with ordinal or categorical data [35]. We based our interpretations of goodness of model fit on cut-off values previously suggested by Brown [33; Table 3]. We also conduct and report χ^2^-tests for both CFA models. However, this test is known to be strongly dependent on the sample size, leading to statistical significance for arbitrarily small deviations of the model-implied and the observed correlation matrices in large samples [33]. Additionally, we conduct a χ^2^-difference-test to investigate whether the two-factor solution fits the data significantly better. Due to the ordinal nature of the data, the scaled version of the χ^2^- difference-test was used [36]. Any potential inflation of the used fit indices based on our very large sample [37, 38], would be inconsequential for our main interest of comparing fit between different models as they are fitted with the same sample size. Still, to rule out any distortions of our results based on the sample size we conducted m-out-of-n Bootstrapping to investigate the likely magnitude of the fit indices if we had had a smaller sample size. The results supported the findings reported in the main text. A comprehensive description and results are provided in Supplement A.

### Measurement Invariance

We tested for measurement invariance by conducting multi-group CFA in four subsamples based on age crossed with gender and in seven subsamples based on medical indication. For both measurement invariance analyses, we adopted the same procedure. First, we calculated the two-factor model CFA for each of the resulting groups, evaluating if this model is a good fit within that group. Second, we conducted multi-group CFA, with several severities of constraints. Model one (M1) was the least constrained model realizing equal factor structure (configural invariance), i.e., the same number of factors and the same assignment of items to factors in all groups. Model two (M2) was more constrained, realizing equal factor loadings (metric invariance), i.e., the same constraints as M1 and the additional constraint that all items have the same loadings on their respective factor in each group.

Model three (M3) was further constrained, realizing equal indicator thresholds (scalar invariance), i.e., the same constraints as M2 and the additional constraint that thresholds between the ordinal levels of the items are equal across groups. Model four (M4) was the most constrained model, realizing equal residuals (strict invariance), i.e., the same constraints as M3 and the additional constraint that the variances of the error terms are equal across groups. As for M3, the used software package lavaan could not return the fit indices according to the robust calculations [34], instead returning NAs. Therefore, we supplemented the scaled version of the fit indices for this analysis step, as suggested by lavaan contributors [39].

The generally advised procedure for multi-group CFA is to fit the models in order of increasing constraint and analyze in each step whether the more constrained model fits worse than the previous one [33]. If the fit for the more constrained model is not worse, invariance at this level is assumed and the analysis continues with the next model of higher constraint. If the fit for the more constrained model is worse, we conduct cumulative multivariate score tests to assess which parameters could be released to increase model fit and achieve partial invariance. In line with our general CFA described above, we primarily based our interpretation of results on fit indices instead of *p*-values and nominal statistical significance. For the multi-group analysis, the critical metric is the difference in these indices between models of increasing constraint. Putnick and Bornstein [40] review criteria proposed in the literature to evaluate whether the difference in fit indices indicates an actual difference in goodness of fit. We chose to apply the most conservative criteria mentioned in that review for each of our fit indices. However, no criteria were mentioned for the TLI. Therefore, we interpret TLI in line with CFI, based on the idea that as we use the same overall criteria for goodness of fit for these indices, we should also use the same criteria for difference of fit. The chosen criteria were: ΔRMSEA < .015; ΔSRMR < .015; ΔCFI < -.002; ΔTLI < -.002.

### Convergent Validity

To assess convergent validity, we analyzed the association between patients’ PHQ-4 scores and other patient reported outcomes, which are expected to correlate with depression and anxiety. For this purpose, we used ordinal logistic regression [specifically proportional odds logistic regression; 41] analysis with the EQ-5D-5L Index, the WAI Score, the work- impairment item of the SIMBO-C, and patients age as predictors, separately for the PHQ-2 and GAD-2 subscales. Prior to the ordinal logistic regression, the predictor variables were standardized to ease interpretation and comparison of the results. The proportional odds assumption and the assumption of absence of collinearity were checked. Likelihood-Ratio χ2- tests were used to evaluate the overall fit of the models against a baseline with only the intercept term and to evaluate the unique contributions of each individual predictor. However, the focus of our interpretations is again not on null hypothesis significance testing but on the explained variance expressed as Nagelkerke *R*^2^ and the effect expressed as Odds Ratios for the standardized predictors. The ordinal logistic regression analysis was chosen to address the ordinal nature of the outcome variable and to be able to estimate effects of each predictor while accounting for the presence of the others. For this analysis we could not utilize the full data set as data for the scores besides PHQ-4 were not available for all patients. The regression analysis still includes a substantial sample of 27,300 patients. The analysis was conducted using the MASS version 7.3-60 [42] and car version 3.1-2 [43] R packages.

Additionally, we conducted the same ordinal logistic regression analysis for the PHQ- 4 composite score (Supplement B). Furthermore, we calculated rank correlation of the PHQ-4 composite score with the scores of the other questionnaires and theoretically indicated single items of questionnaires (Supplement C). Those results were in line with the results of the ordinal regression analyses reported in the main text.

## Results

### Sample characteristics

The sample consisted of 66,412 patients; 29,010 (43.7%) were female. The mean age was 59.6 (*SD* 13.3) years; age ranged from 18 to 98, with a median of 59. Patients were treated for the following medical indications: orthopedic (*n* = 22,676), cardiac (*n* = 19,604), metabolic (*n* = 8,429), respiratory (*n* = 7,662), neurological (*n* = 3,762), gastrointestinal (*n* = 1,749), oncological (*n* = 1,606) and various smaller indications (*n* = 924).

For all 66,412 patients PHQ-4 data, age, and gender were available. Data for the EQ-5D-5L, WAI, and SIMBO-C items and scores were available for subsets of the full sample (see Table 4 for subsample sizes). According to a criterion of ≥ 6 for the composite score, 14.0% of the patients reported at least moderate depression and/or anxiety symptoms. According to a criterion of ≥ 3 for the PHQ-2 and GAD-2 sub-scores, 18.6% of the patients had depression symptoms and 16.3% had anxiety symptoms.

### Item Characteristics and Internal Consistency

The mean PHQ-4 composite score in the sample was 2.2 (*SD* = 2.7). The mean PHQ-2 depression score was 1.2 (*SD* = 1.5), while the mean GAD-2 anxiety score was 1.1 (*SD* = 1.4). All score and subscore means and *SD*s can be found in Table 2.

**Table 2.** Mean Scores for Full Sample and Subsamples.

| | | Mean ($SD$) | | | N (%) |
| --- | --- | --- | --- | --- | --- |
|  |  | Composite (PHQ-4) | Depression (PHQ-2) | Anxiety (GAD-2) |  |
| Full Sample |  | 2.2 (2.7) | 1.2 (1.5) | 1.1 (1.4) | 66,412 (100%) |
| Female | < 60 years | 3.1 (3.0) | 1.5 (1.6) | 1.5 (1.6) | 14,511 (21.8%) |
| | $\geq 60$ years | 2.3 (2.6) | 1.1 (1.4) | 1.1 (1.4) | 14,499 (21.8%) |
| Male | < 60 years | 2.2 (2.7) | 1.2 (1.5) | 1.0 (1.4) | 19,587 (29.5%) |
| | $\geq 60$ years | 1.6 (2.2) | 0.9 (1.3) | 0.7 (1.2) | 17,815 (26.9%) |

The item intercorrelations were: *r*_PHQ-2.1, PHQ-2.2_ = .739; *r*_PHQ-2.1, GAD-2.1_ = .613; *r*_PHQ-2.1, GAD-2.2_ = .621; *r*_PHQ-2.2, GAD-2.1_ = .719; *r* _PHQ-2.2, GAD-2.2_ = .737; *r*_GAD-2.1, GAD-2.2_ = .732, showing somewhat higher-correlations between items within subscale than across subscales. The correlation between the PHQ-2 and GAD-2 subscores was high at *r* = .744, but below typical cut-offs for poor discriminant validity [r = .85; 33]. Item-total correlations were PHQ-2-1: .877, PHQ-2.2: .923, GAD-2.1: .985, GAD-2.2: .987. The internal consistency of PHQ-2 and GAD-2 as measured with *Mc Donald’s* ω were ω = .775 and ω = .758, respectively, while the internal consistency of the PHQ-4 was ω = .852. Figure 1 depicts an overview of response options, scores and their associations.

**Figure 1.**
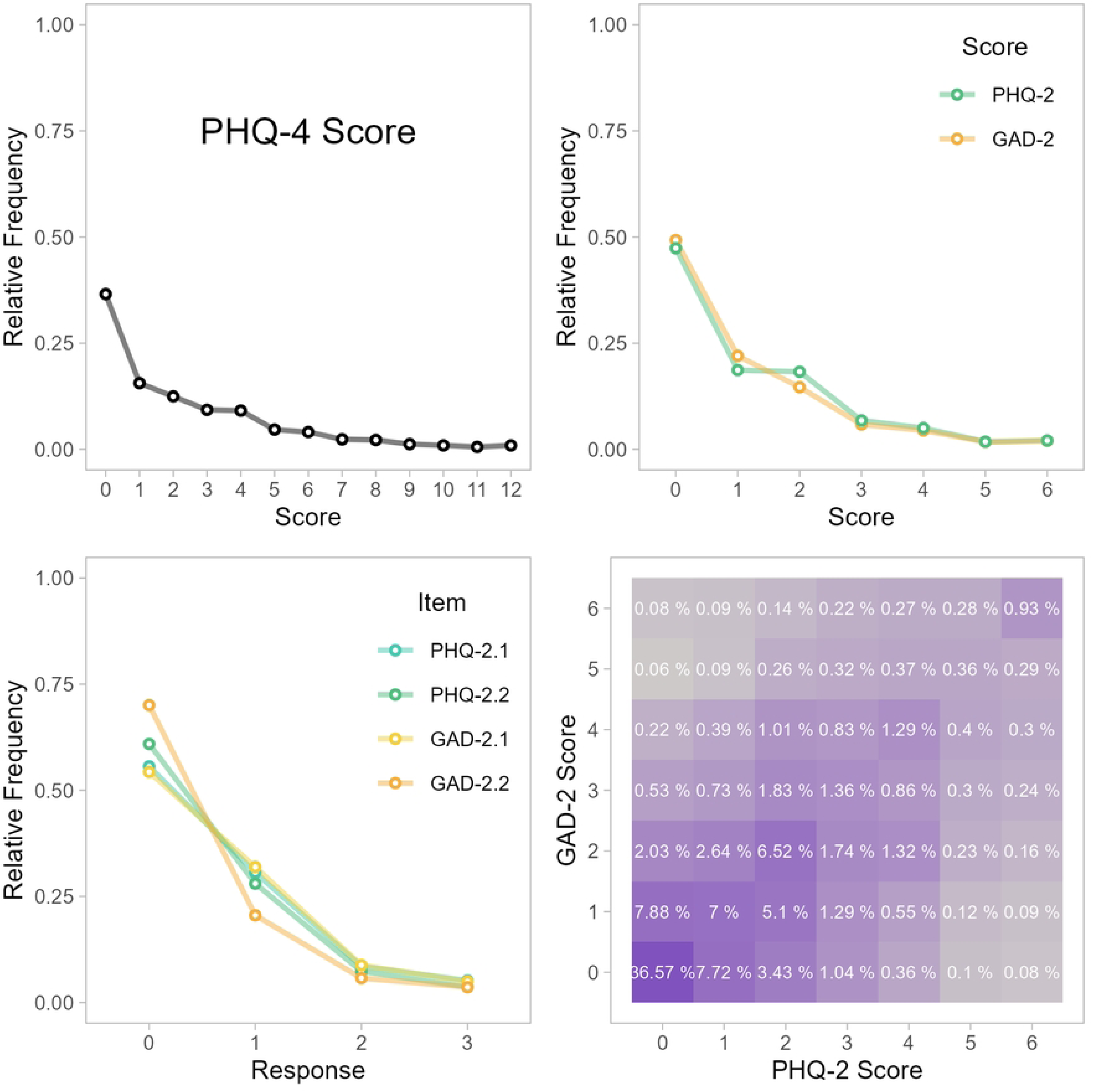
Distribution of PHQ-4 Data. *Note.* The top left, top right, and bottom left panels depict the relative frequencies of response options and scores in the sample for the PHQ-4 composite score, the subscale scores, and the individual items respectively. The bottom right panel depicts the association between the PHQ-2 and the GAD-2 subscales as relative co-occurrence frequencies of subscale scores.

### Confirmatory factor analysis

To evaluate the factorial validity of the PHQ-4, we conducted confirmatory factor analysis (CFA). The four chosen fit indices all showed a good fit for the two-factor solution (RMSEA [90% *CI*] = .014 [.006; .025], SRMR = .001, CFI > .999, TLI > .999), while only two of four (SRMR and CFI) showed a good fit for the one-factor model (RMSEA (90% *CI*) = .171 [.164; .178], SRMR = .024, CFI = .977, TLI = .931). The χ^2^-tests for both the two-sample (χ^2^(1) = 7.046, *p* = .008) and one-sample (χ^2^(2) = 1991.96, *p* < .001) model were statistically significant. The statistically significant *p*-values for the χ^2^-tests for both CFA models would usually suggest that neither have a good fit. However, this test is known to be strongly dependent on the sample size, leading to statistical significance for arbitrarily small deviations of the model-implied and the observed correlation matrices for large samples [33]. The results of the χ^2^-difference-test supported a statistically significant better fit for the two-factor model (χ^2^ (1) = 1587, *p* < .001); but again, this analysis is influenced by sample size. Figure 2 shows the path diagrams for the one- and two-factor solutions.

**Figure 2.**
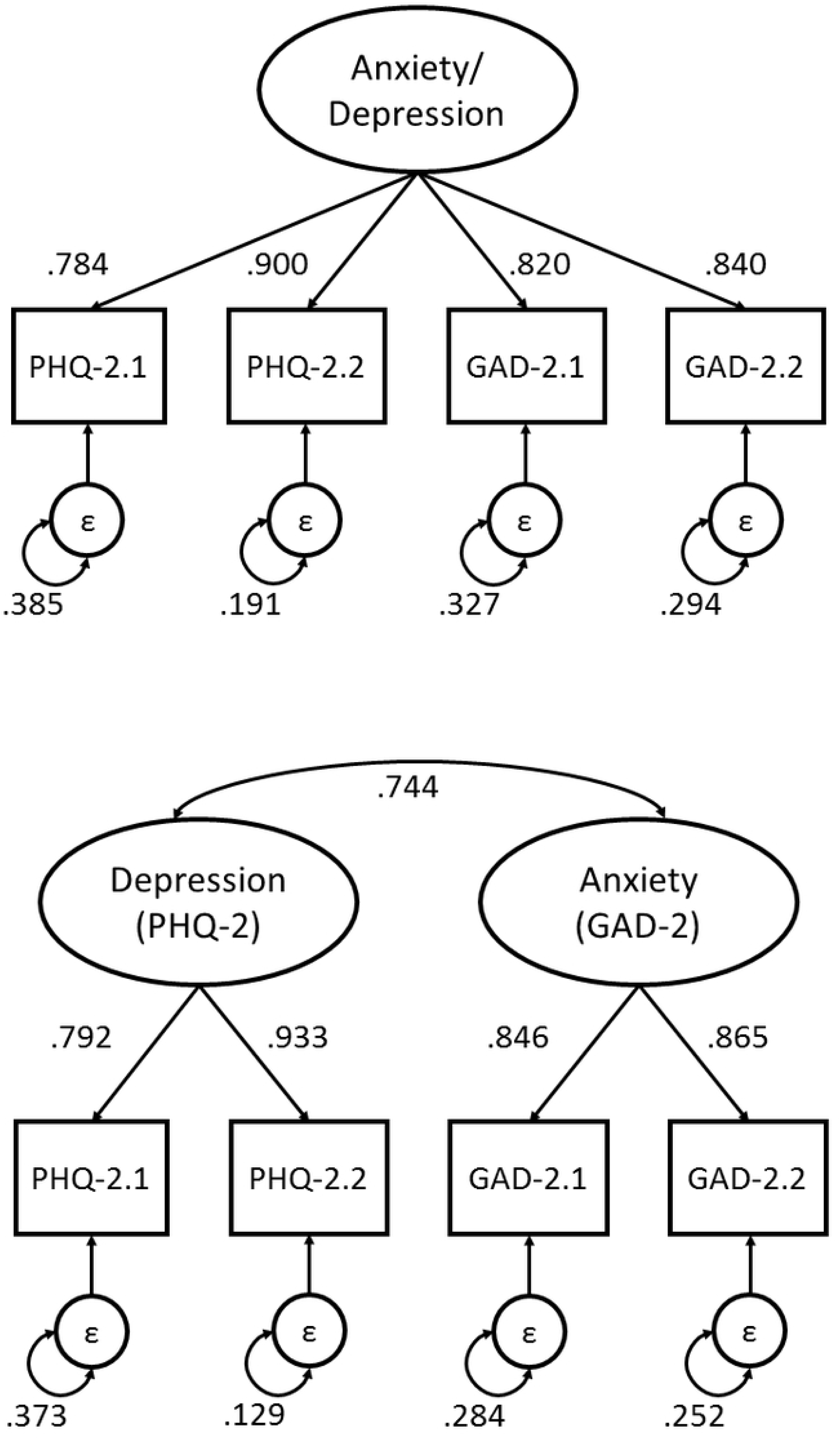
Path Diagrams for the One- and Two-factor Solutions of the PHQ-4. *Note.* Depicted are factor loadings, error variances, and factor covariance of the standardized solutions.

### Measurement Invariance – Age and Gender

To achieve similar group sizes, we grouped by gender and by the median age of our sample (i.e., 59 years). Descriptive statistics and sample sizes of the four resulting groups can be found in Table 2. The results for the two-factor CFA within each group can be obtained from Table 3. Overall, all four groups showed a relatively good fit, with the group of younger females showing the worst. However, they still showed good fit on three of the four fit indices we used, while also showing clearly better fit for the two-factor solution than the alternative one-factor solution (χ^2^(2) = 623.11, *p* < .001; RMSEA [90% *CI*] = .188 [.175; .201], SRMR = .028, CFI = .971, TLI = .913).

**Table 3.**
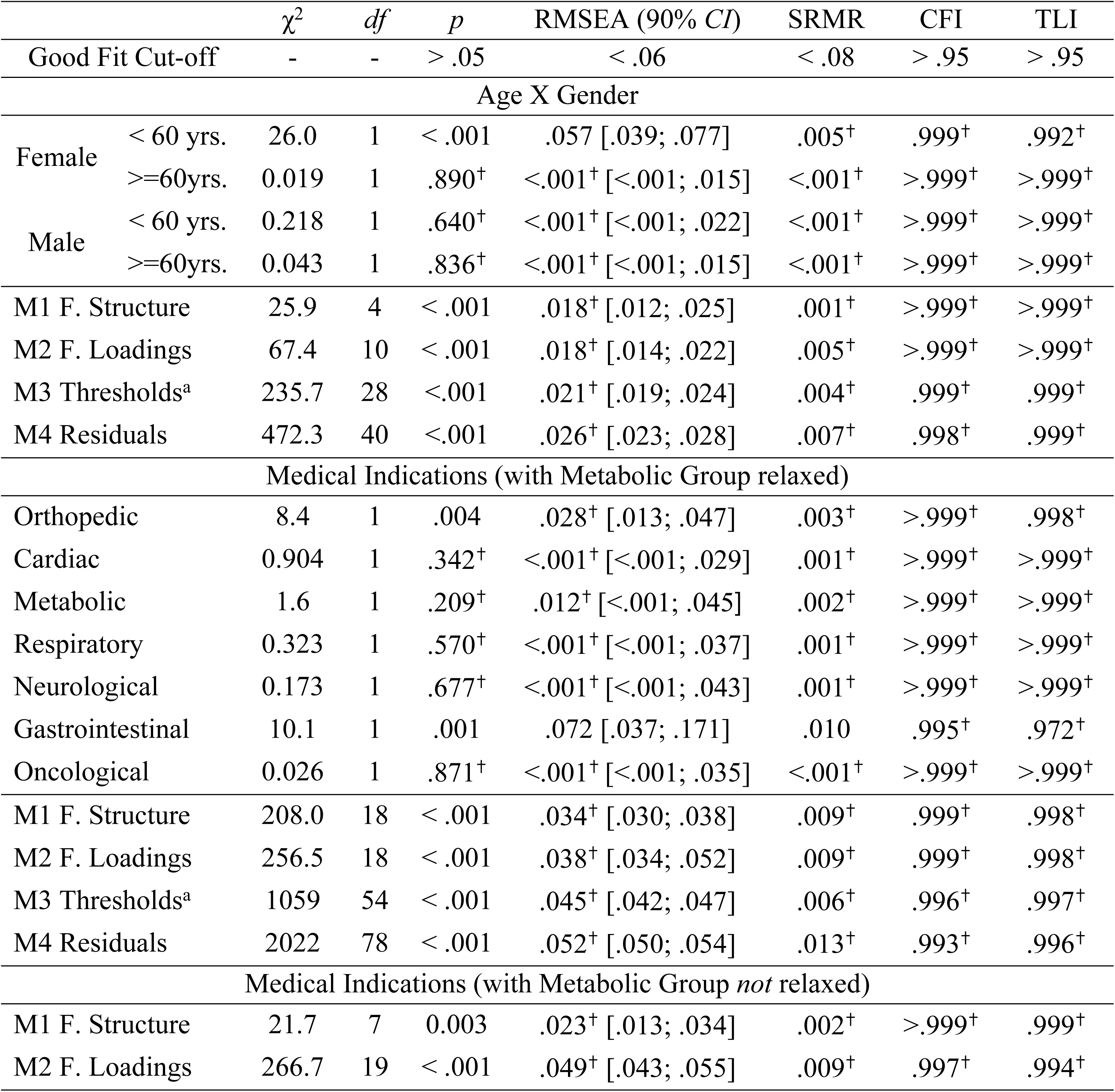
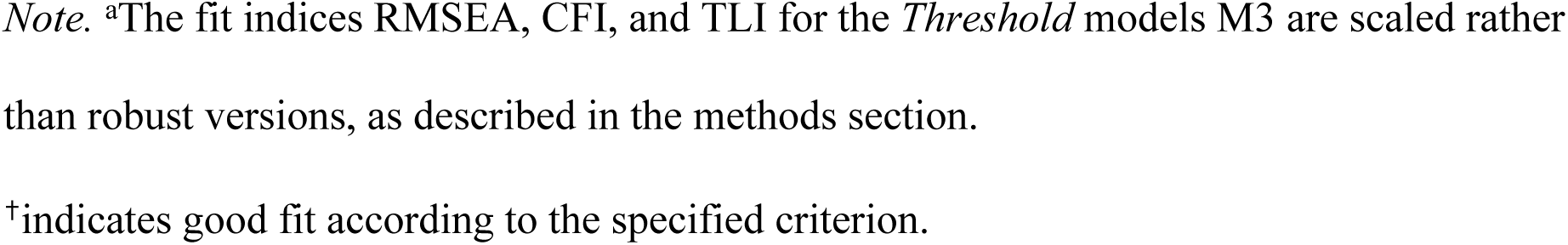
Measurement Invariance: Model Fit for Subgroups and Constraint Models.

Next, we conducted the multi-group CFA for M1 through M4. All models showed good fit according to all four fit indices (see Table 3). According to our criteria of difference in fit, M2 did not differ in fit from M1, M3 did not differ from M2, and M4 did not differ from M3 (see Table 4). Therefore, we can assume strict invariance over the tested age and gender groups.

**Table 4.** Measurement Invariance: Difference in Fit for Constraint Models.

| | $\Delta\chi^2$ | $\Delta df$ | $p$ | $\Delta RMSEA$ | $\Delta SRMR$ | $\Delta CFI$ | $\Delta TLI$ |
| --- | --- | --- | --- | --- | --- | --- | --- |
| $\Delta$ Fit Cut-off | - | - | > .05 | < .015 | < .015 | > -.002 | > -.002 |
| Age X Gender |  |  |  |  |  |  |  |
| M2 – M1 | 39.46 | 6 | < .001 | <.001<sup>†</sup> | .004<sup>†</sup> | <-.001<sup>†</sup> | <-.001<sup>†</sup> |
| M3 – M2 | 216.7 | 18 | < .001 | .003<sup>†</sup> | .001<sup>†</sup> | -.001<sup>†</sup> | <-.001<sup>†</sup> |
| M4 – M3 | 237.6 | 12 | < .001 | .004<sup>†</sup> | .003<sup>†</sup> | -.001<sup>†</sup> | <-.001<sup>†</sup> |
| Medical Indications (with Metabolic Group relaxed) |  |  |  |  |  |  |  |
| M2 – M1 | -5.43 | 0 | < .001 | .004<sup>†</sup> | <.001<sup>†</sup> | <-.001<sup>†</sup> | <-.001 |
| M3 – M2 | 991.6 | 36 | < .001 | .007<sup>†</sup> | -.003<sup>†</sup> | -.003 | -.001<sup>†</sup> |
| M4 – M3 | 970.4 | 24 | < .001 | .007<sup>†</sup> | .007<sup>†</sup> | -.003 | -.001<sup>†</sup> |
| Medical Indications (with Metabolic Group *not* relaxed) |  |  |  |  |  |  |  |
| M2 – M1 | 214.1 | 12 | < .001 | .022 | .007<sup>†</sup> | = -.001<sup>†</sup> | -.002 |

*Note:* <sup>†</sup>indicates good fit according to the specified criterion.

### Measurement Invariance – Medical Indications

For this analysis, we included the seven largest medical indications of the sample, totaling 65,488 patients; 98.6% of our total sample. Overall, all seven groups showed a relatively good fit to the two-factor model (see Table 3), with the group of gastrointestinal patients showing the worst fit. However, they still showed good fit on two of the four fit indices we used. For this group the one-factor solution was not an ideal fit either (χ^2^(2) = 31.8, *p* < .001; RMSEA [90% *CI*] = .137 [.098; .180], SRMR = .020, CFI = .985, TLI = .954), performing worse on all indices (Δχ^2^(1) = 20.21, *p* < .001; ΔRMSEA [90% *CI*] = .029; ΔSRMR = .01, ΔCFI = -.011, ΔTLI = -.018), reaching our threshold for worse fit for the RMSEA, CFI, and TLI, but not the SRMR. Therefore, our overall results for the group of gastrointestinal patients supports the two-factor structure, although not as robustly as for the other groups.

Next, we conducted the multi-group CFA for M1 through M4. In our initial analysis, we found only moderate support for the more constraint model M2 fitting as well as M1, with two of our four fit indices exceeding our cut-off for equal fit (Δχ^2^(12) = 214.1, *p* < .001; ΔRMSEA [90% *CI*] = .022; ΔSRMR = .007, ΔCFI = -.001, ΔTLI = -.002). We used multivariate score tests (i.e.; the Lagrange Multiplier test) implemented in lavaan to identify the most impactful possible changes to the model. This suggested that the strongest effect was given by releasing the constraint that the factor loadings for the anxiety factor for the metabolic group were equal to those of the other groups (χ^2^(1) = 63.794, *p* = <.001). We therefore recalculated the analysis of invariance over medical indication, with a modified two-factor model which constraints the factor loadings to be equal except for all groups, except the metabolic one. The results of this analysis showed good fit for M1 through M4 according to all four fit indices (see Table 3). M2 did not fit worse than M1. The differences in fit indices between M2 and M3 as well as between M3 and M4 only exceeded our very strict cutoffs for one fit index by a very narrow margin. Overall, we interpret these results as supporting strict invariance for all tested medical indications, except for the metabolic group. For that subgroup, we could only support the presence of configural invariance.

### Convergent Validity

The conducted ordinal logistic regression showed that all used predictors exhibited expected associations with the PHQ-2 and GAD-2 subscores. The overall models explained 24.5% of the variance in the PHQ-2 subscore (*R*^2^ = .245, χ^2^(4) = 7268.69, *p* = <.001), and 20.4% of the variance in the GAD-2 subscore (*R*^2^ = .204, χ^2^(4) = 5865.16, *p* = <.001). Detailed model parameter can be found in Table 5.

**Table 5.** Convergent Validity: Ordinal Logistic Regression Parameters.

| | $\beta$ | $\chi^2$ | $df$ | $p$ | $R^2$ | OR (95% CI) |
| --- | --- | --- | --- | --- | --- | --- |
| PHQ-2 Subscore |  |  |  |  |  |  |
| Full Model | - | 7264.69 | 4 | < .001 | 0.245 | - |
| Age | -0.193 | 2060.39 | 1 | < .001 | - | 0.825 [0.806; 0.843] |
| EQ-5D-5L | -0.598 | 59.88 | 1 | < .001 | - | 0.550 [0.536; 0.564] |
| WAI | -0.676 | 1513.78 | 1 | < .001 | - | 0.509 [0.491; 0.527] |
| SIMBO-C Impairment | -0.128 | 283.55 | 1 | < .001 | - | 0.880 [0.852; 0.909] |
| GAD-2 Subscore |  |  |  |  |  |  |
| Full Model | - | 5865.16 | 4 | < .001 | 0.204 | - |
| Age | -0.156 | 183.09 | 1 | < .001 | - | 0.855 [0.836; 0.875] |
| EQ-5D-5L | -0.561 | 1830.53 | 1 | < .001 | - | 0.570 [0.556; 0.585] |
| WAI | -0.649 | 1369.24 | 1 | < .001 | - | 0.523 [0.505; 0.541] |
| SIMBO-C Impairment | -0.226 | 182.78 | 1 | < .001 | - | 0.798 [0.772; 0.825] |

*Note:* $\chi^2$ refers to Likelihood Ratio tests.

For the PHQ-2 model, for every one standard deviation increase in age the odds of observing a higher level of the PHQ-2 decreased by 17.54%, for every one standard deviation increase in the EQ-5D-5L Index the odds decreased by 45.02%, for every one standard deviation increase in the WAI-Score the odds decreased by 49.13%, and for every one standard deviation increase in the SIMBO-C work impairment item the odds decreased by 11.98%; holding all other variables constant. For the GAD-2 model, for every one standard deviation increase in age the odds of observing a higher level of the GAD-2 decreased by 14.47%, for every one standard deviation increase in the EQ-5D-5L Index the odds decreased by 42.96%, for every one standard deviation increase in the WAI-Score the odds decreased by 47.73%, and for every one standard deviation increase in the SIMBO-C work impairment item the odds decreased by 20.19%; holding all other variables constant.

The assumptions of proportional odds and no collinearity were checked and found to be largely fullfilled, with only questionable values for the proportionallity of odds for the SIMBO-C item for the highest levels of the PHQ-2 (see Supplement B).

## Discussion

Our findings offer support for the PHQ-4 as a valid and reliable screening tool for the identification of anxiety and depression within the context of rehabilitation settings. Utilizing confirmatory factor analysis (CFA), it could be determined that the two-factor structure of the PHQ-4, distinguishing between anxiety and depression, holds in the rehabilitation setting, supporting its factorial validity. Furthermore, as the multi-group CFA showed, this structure also holds for age and gender based sub-groups, suggesting that the PHQ-4 is applicable for a wide range of rehabilitation patients. Correlational analysis supported convergent validity of the PHQ-4, using existing measures of health, function, impairment and work ability. Finally, while the reliabilities of the GAD-2 and PHQ-2 subscales were acceptable, the reliability of the PHQ-4 composite score was better. These findings align with previous validation studies of the PHQ-4 for general and primary care populations [6].

For the confirmatory factor analysis, we compared two potential models: a one-factor model suggesting that the PHQ-4 measures only a single dimension of psychological distress and a two-factor model suggesting that the PHQ-4 measures anxiety and depression. While the one-factor model showed not ideal fit for two out of the four chosen fit indices, the two-factor model showed excellent fit for all four. In line with previous validations of the PHQ-4 [5, 18–21, 44], this supported the existence of two underlying dimensions of the PHQ-4, as intended by psychological theory.

Our analysis of measurement invariance supported strict invariance for the PHQ-4 over the tested age and gender groups as well as for six out of seven medical indications. For the group of patients in metabolic rehabilitation we could only show configural invariance. Future studies employing comparative analyses of sensitivity and specificity of the PHQ-4 between different medical indications could illuminate whether the idiosyncrasy of the metabolic group is only of academic interest or warrants unique cutoff values in screening for anxiety and depression in this group. Overall, our results underline the suitability of the PHQ- 4 as a reliable tool for routine psychological assessment, particularly within a heterogeneous rehabilitation population, with the mentioned limitation for metabolic patients.

Our analysis of convergent validity demonstrated that the PHQ-4 showed expected patterns of associations with other measures routinely used in the rehabilitation context. This further supports the notion that the PHQ-4 operates as intended in this population. Our ordinal logistic regression clearly demonstrated associations of the PHQ-4 subscores with age, work ability, impairment, and quality of life. The results of our correlational analysis reported in the supplement further showed that the PHQ-4 mainly exhibited medium to strong correlations in the expected directions with theoretically implicated scores and items from existing instruments measuring self-rated health and function, workability / work impairment, and perceived mental resources. Furthermore, as reported before [45], we found higher levels of self-reported anxiety and depression in women. Age showed a trivial-to-small negative association with the PHQ-4 score in our sample. Previous studies have reported mixed results on the association of depression with age. While some studied samples show positive and some negative correlations, this can be reconciled by assuming an inverted U-shape of depression and anxiety over age [46–49]. It is however important to note that our results may have been influenced by the predominance of older individuals in our sample (median age of 59).

Notably, the prevalence of probable depression (18.6%) and anxiety (16.3%) in our rehabilitation sample exceeded rates observed in the German general population [with 10.4% and 9.8%, respectively; 50]. This is in line with prior research suggesting that chronic diseases and worse self-rated health contribute to heightened psychological distress [50–55]. Given the known impact of depression and anxiety on rehabilitation outcomes [10–15], our findings emphasize the importance of routine mental health screening in rehabilitation settings. This practice is essential not only for improving patient outcomes but also for enabling healthcare providers to use resources more efficiently, optimize treatment strategies, and ultimately facilitate improved rehabilitation results.

Overall, our findings demonstrate that the PHQ-4 is appropriate for screening for anxiety and depression in rehabilitation patients despite the unique psychological challenges faced by this heterogeneous patient collective. This study provides firm evidence that this holds for Austrian rehabilitation patients. However, we believe that given the current study’s substantial sample size and the corroborating findings from the subgroup analysis, it is likely that the study’s findings will generalize to rehabilitation populations in various countries. Still, empirical studies in these populations are necessary to provide conclusive evidence for this interpretation.

### Strength and Limitations

The strengths of the study include the large sample size, the diversity of the 15 included rehabilitation centers, and the use of robust statistical methods. The current study applied confirmatory factor analysis with scale and distribution appropriate estimators and properly adjusted fit indices, as compared to many previous studies applying techniques not appropriate for the scale of the data, such as Maximum Likelihood estimation or not ideal to confirm factorial validity such as exploratory factor analysis (EFA) or principal component analysis (PCA).

The main limitation of this study is the absence of clinical interviews precluded an analysis of sensitivity and specificity, which would further establish its diagnostic accuracy. Additionally, the 4-item composition of the PHQ-4 doesn’t allow for more complex statistical models, such as second-order or bifactor structures, due to identification constraints, limiting us to the comparison of one- and two-factor solutions.

## Data Availability

The dataset of this study contains sensitive patient information and cannot be shared publicly. Ethical restrictions have been imposed by the data protection officials of the Pensionsversicherung, who may be contacted for data access. Inquiries should be directed to: Datenschutzbeauftragter, Friedrich-Hillegeist-Straße 1, 1020 Wien

